# Alleviating negative symptoms in schizophrenia using a Virtual Reality-based therapy targeting social reward learning (ENGAGE): Protocol for a randomised, controlled, assessor-blind pilot study

**DOI:** 10.1101/2025.08.20.25334133

**Authors:** Valentin M. Donath, Emma Slebsager Ries, Lise Mariegaard, Tina Dam Kristensen, Bjørn H. Ebdrup, Gregory P. Strauss, Patrick M. Fisher, Egill Rostrup, Merete Nordentoft, Rikke Hilker, Marianne Melau, Claudi L. Bockting, Nikolai Albert, Martine J. van Bennekom, Karen S. Ambrosen, Louise Birkedal Glenthøj

## Abstract

In individuals with schizophrenia spectrum disorders (SSD), negative symptoms (NS) are known to be associated with low quality of life, predictive of adverse long-term outcomes, and barriers to relevant life goals such as educational, vocational, and social attainment. As social cognition, processes of reward appraisal, and anticipation are impaired in individuals with SSD, these dysfunctions are likely to be intertwined with the pathogenesis of NS. Despite their debilitating nature, there remains a scarcity of treatment options for NS, as they, unlike positive symptoms, are largely unaffected by pharmacological interventions. Among indications that psychosocial interventions can reduce NS, more robust evidence is warranted for interventions that directly target NS. In light of the recent advances in Virtual Reality-assisted psychotherapy (VRT) for the treatment of positive symptoms in schizophrenia (i.e., paranoia and auditory hallucinations), this randomised, assessor-blind, controlled pilot study sets out to test the feasibility and acceptability of a novel VRT aimed at alleviating NS through targeting social reward learning (ClinicalTrials.gov registration ID: NCT06993831). The study will enrol 30 outpatients from the public mental health services of Greater Copenhagen, Denmark, who will be equally randomised to receive either treatment as usual (TAU) or TAU combined with 10 sessions of individual, VR-assisted psychotherapy. Feasibility and acceptability endpoints will be supplemented by clinical interviews and ecological momentary assessments (EMA) for indications of treatment efficacy regarding positive and negative symptomatology, functional outcome, and quality of life. Additionally, neurobiological and behavioural correlates of the intervention will be explored by magnetic resonance imaging (MRI).

## Introduction

Schizophrenia is a severe mental illness associated with extremely high rates of functional disability (1), global disease burden (2,3), healthcare utilisation costs (4,5), and premature mortality (6). Negative symptoms (NS), commonly conceptualised as anhedonia, avolition, asociality, alogia, and blunted affect, are strong predictors of many of the poor clinical and functional outcomes associated with schizophrenia (7). While positive symptoms can be effectively targeted by antipsychotic medication (8), robust evidence for treatments directly aimed at alleviating NS is lacking (9,10). Even within extant specialised early intervention programmes, approximately half of the treated patients will not experience a reduction in NS (11). In light of their considerable impact on functional long-term outcomes (12), effective treatments for NS have been identified as a critical unmet need in schizophrenia therapeutics (13).

Abnormalities in numerous aspects of reward processing (e.g., reward anticipation, reinforcement learning, effort-cost computation, hedonic reactivity, value representation) governed by cortico-striatal circuitry have been consistently associated with NS, particularly anhedonia and avolition (14–17). Other psychological mechanisms also predict elevations in specific NS domains, including defeatist performance beliefs, asocial beliefs, low pleasure beliefs, and low expectancies for success (18). Impairments in several domains of social cognition have also been linked to asociality and NS more broadly (19–21). These psychological and neural processes reflect viable mechanisms of action that could be targeted in interventions for NS.

Initial evidence suggests that psychosocial interventions can improve negative symptoms via approaches utilising Cognitive Behaviour Therapy, Social Skills Training, Cognitive Remediation, Social Cognition Training, Behavioural Activation, and Positive Affect Training (22–24). These interventions are believed to act on NS through their effect on putative mechanisms, such as reward processing, social cognition, and dysfunctional beliefs (25–28). However, given that the magnitude of improvement is typically at a small effect size (29,30), there is considerable need to amplify the effects of psychosocial treatments to yield clinically meaningful changes capable of changing real-life functioning.

Virtual reality-based therapy (VRT) is a therapeutic tool that holds promise for amplifying the effects of standard psychosocial approaches (31,32). VRT requires patients to immerse themselves in simulated, realistic surroundings that depict every-day interactions with objects and people in common settings. It has proven safe and efficacious for treating positive symptoms in schizophrenia (33–35), outperforming many other nonpharmacological treatments in this domain (36). Emerging evidence also suggests beneficial effects of VRT on social functioning (37). However, findings regarding their impact on NS are mixed (38). Inconsistent effects in past studies may reflect differences in the mechanisms of action that were targeted. Specifically, the VR interventions in previous studies have primarily been designed to improve Theory of Mind (39), and social skills (40), which may not have comprehensively targeted the range of mechanisms most relevant to negative symptoms. Thus, there remains a critical need to investigate VR-therapies specifically developed to improve NS by targeting the most critical mechanisms (e.g., reward processing, dysfunctional beliefs, multiple social cognitive processes).

### Hypotheses

The present pilot study aims to test the feasibility and acceptability of a novel VR-based psychotherapeutic intervention for patients with an SSD with NS. We hypothesise that VR assisted psychotherapy for NS patients

1. is feasible and acceptable, and
2. will prove superior to treatment as usual (TAU) in reducing NS and improving daily functioning, signalling treatment efficacy.

## Methods

### Study design

The study is sponsored by and conducted at Mental Health Centre Copenhagen, Mental Health Services, Capital Region of Denmark. In an assessor-blind, mixed-methods, randomised controlled trial (RCT), 30 participants with an SSD diagnosis (ICD-10 F2x) will be recruited from specialised outpatient care facilities for patients with SSD (so-called OPUS and F-ACT teams) within the Copenhagen Mental Health Services. Following an eligibility screening, participants will be randomly assigned to one of two study arms:

1. Treatment as Usual (TAU; control group) or
2. TAU plus 10 sessions of VR-assisted psychotherapy (intervention group).

Clinical assessments, MRI, and data from seven-day Ecological Momentary Assessment (EMA, cf. “Secondary outcomes”) epochs will be obtained at baseline and upon treatment completion 14 weeks after baseline (see Fig. 1).

**Fig 1.**
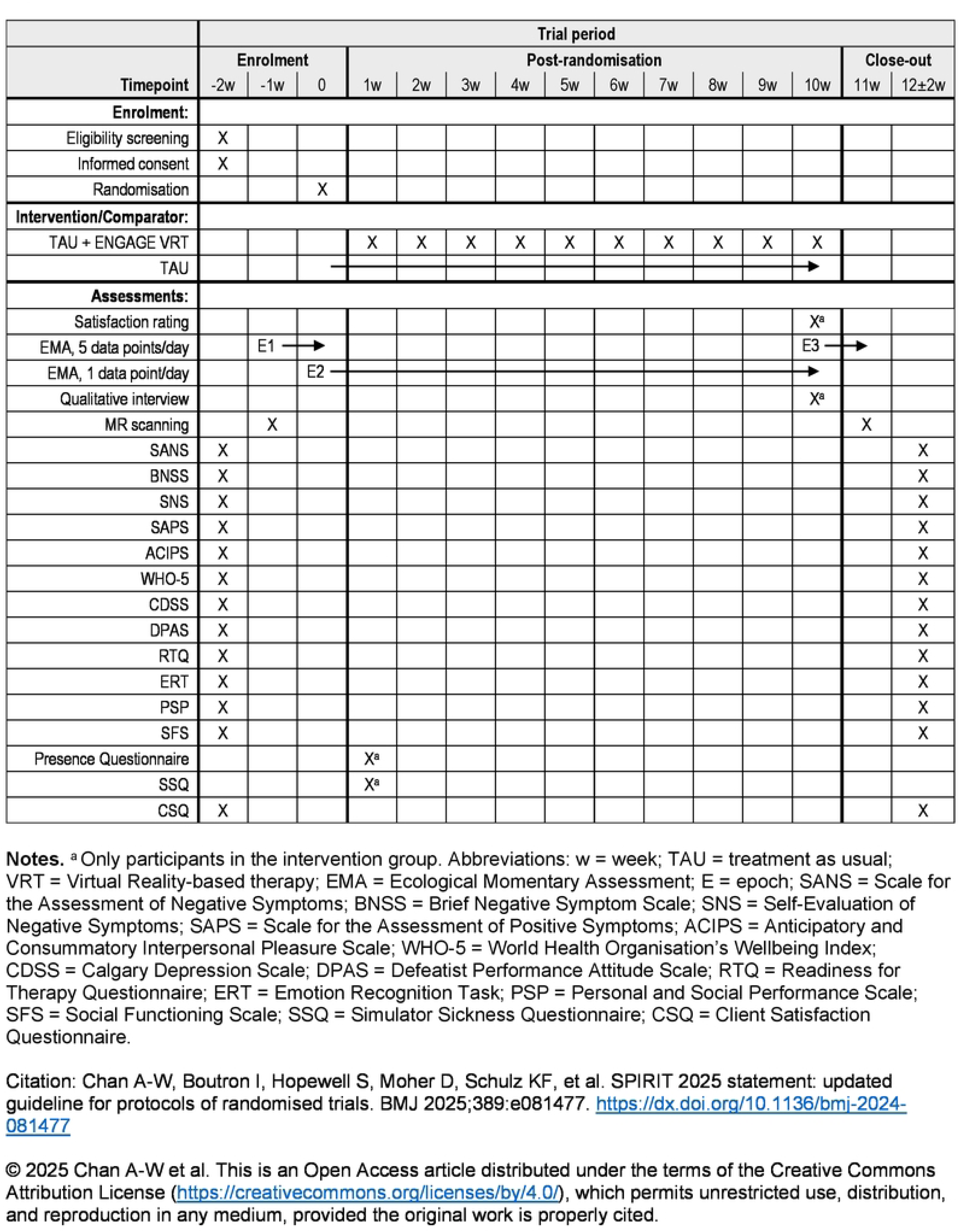
SPIRIT schedule of enrolment, interventions, and assessments.

The study is registered at ClinicalTrials.gov (NCT06993831). See Appendix S1 for the Standard Protocol Items: Recommendations for Interventional Trials (SPIRIT) checklist, and Appendix S2 for the trial protocol approved by the National Committee on Health Research Ethics for the Capital Region of Denmark.

### Participants

The sample size of the present feasibility study is set at *n* = 30 in line with recommendations for pilot studies (41,42). Patients fulfilling the criteria presented in Table 1 will be admissible to the study.

**Table 1.**
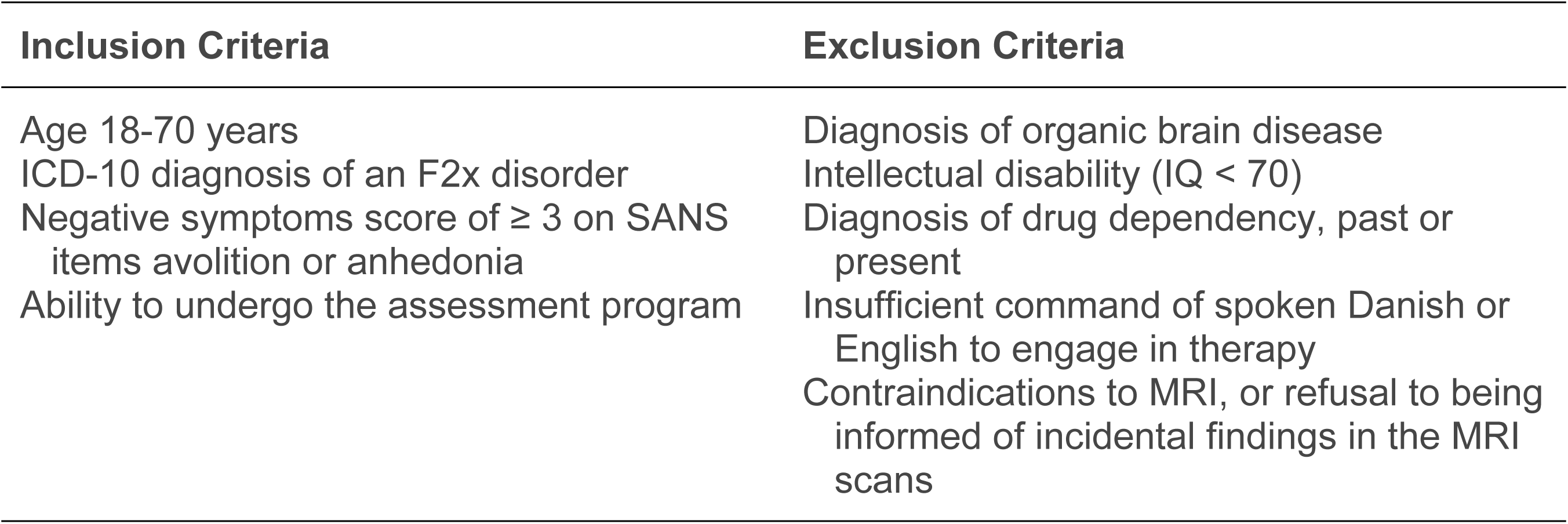
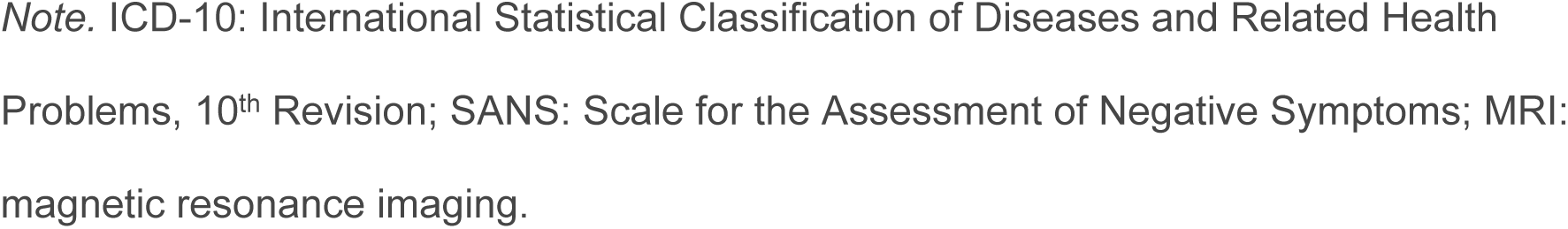
Inclusion and exclusion criteria for study participants.

*Inclusion and exclusion criteria for study participants*
| **Inclusion Criteria** | **Exclusion Criteria** |
| --- | --- |
| Age 18-70 years<br>ICD-10 diagnosis of an F2x disorder<br>Negative symptoms score of $\geq 3$ on SANS items avolition or anhedonia<br>Ability to undergo the assessment program | Diagnosis of organic brain disease<br>Intellectual disability (IQ < 70)<br>Diagnosis of drug dependency, past or present<br>Insufficient command of spoken Danish or English to engage in therapy<br>Contraindications to MRI, or refusal to being informed of incidental findings in the MRI scans |

*Note.* ICD-10: International Statistical Classification of Diseases and Related Health Problems, 10<sup>th</sup> Revision; SANS: Scale for the Assessment of Negative Symptoms; MRI: magnetic resonance imaging.

### Setting and procedure

The study is conducted by VIRTU Research Group at Mental Health Centre Copenhagen, Mental Health Services CPH, in collaboration with the Centre for Neuropsychiatric Schizophrenia Research (CNSR) at Mental Health Centre Glostrup, Denmark. MRI is performed at the Functional Imaging Unit (FIU), Rigshospitalet – Glostrup, Denmark.

Clinical, behavioural (EMA), and MRI data will be collected at baseline and at follow-up, 14 weeks after baseline, by a trained psychologist functioning as study assessor blinded to group allocation. Another psychologist will collect qualitative data on treatment acceptability (cf. “Qualitative data”).

### Randomisation and blinding

An unstratified, blocked randomisation sequence will be generated by a third party, uninvolved in the study. Study assessors will be blinded to sequence and block size. Once a participant has completed baseline assessments, unblinded study staff (i.e., therapists and study assistants) will allocate the participant to either study arm utilising the randomisation functionality of the Research Electronic Data Capture system (REDCap) (43).

Study assessors will be blinded to group allocation until after statistical analyses have been completed. Participants, therapists, assistants, and other non-blinded study staff will be instructed not to reveal group allocation to assessors.

### Intervention

In addition to standard care, participants allocated to the intervention group will be offered 10 sessions of manualised, modularised, VR-supported psychotherapy delivered by licensed psychologists with expertise in working with VR-interventions for psychosis. A portion of each session will be spent in VR. Visual and auditory stimuli will be presented using the VR software Social Worlds (44) and VR Moodboost (45), which can be used to emulate a variety of real-life surroundings such as a park, a bus, a home, or a supermarket. Participants can virtually move around in these surroundings or engage in social interactions with avatars, whose voices are produced by real-time distortion of the therapist’s voice. The individually modified virtual reality scenarios are designed to stimulate social reward as operant conditioning principles, motivation and the experience of positive affect in the participant. Alongside the in-session experiences, the participant is given weekly activity tasks to be carried out at home between sessions. The participants are asked to register anticipatory pleasure before performing an in-session or at-home activity, and to register participatory pleasure directly after completing it.

The intervention is divided into four modules: Introduction, Motivation, Social Reward, and an extra module for individual adaptation (see Appendix S3 for a session overview of the ENGAGE VR therapy.). Sessions on attention focus and motivating an avatar are adapted from Meins et al.’s (45) VR-SOAP intervention for social functioning in psychosis.

To bridge sessions and real-life behaviours, the weekly at-home activities will be planned using a study app (46). In the study app, psychoeducational material on NS will be made available to participants in the intervention group after baseline assessment, and to participants in the control group after follow-up assessment. In-app graphs visualising the participant’s anticipatory and consummatory pleasure ratings of the in-session VR tasks and at-home activities will be used therapeutically to illustrate patterns in the participant’s experience of pleasure, such as biased discrepancies between anticipatory and consummatory pleasure, and patterns of development over the course of therapy, such as an increase in anticipatory pleasure.

### Control group (Treatment as usual, TAU)

Participants in either group will continue to receive TAU provided by their respective outpatient clinic of the Copenhagen Mental Health Services. For patients with an SSD, TAU typically consists of supportive counselling with a nurse and regular psychiatric consultations, and may also include sessions with a psychologist or social worker, as well as group or individual therapy. The specific composition and intensity of TAU is determined by the outpatient clinic managing the treatment.

### Primary outcome measures

Feasibility and acceptability will be evaluated as:

1. Recruitment of 80% of the target sample within 15 months.
2. Retention to study protocol of 70% at cessation of therapy (10 sessions).
3. Satisfaction rating of ≥7 on a 10-point Likert scale by 80% of participants in the intervention group.

### Secondary outcome measures

#### Clinical, cognitive, and functional outcomes

To obtain indications of a potential for treatment efficacy, clinical, cognitive, and functional outcomes, as presented in Table 2, will be assessed at baseline and at 14 weeks follow-up.

**Table 2.** Secondary clinical, cognitive and functional outcomes and measures.

| Outcome | Measure | Data Source |
| --- | --- | --- |
| Clinical assessment |  |  |
| Severity of negative symptoms | Scale for the Assessment of Negative Symptoms (SANS, 47) | A |
|  | Brief Negative Symptom Scale (BNSS, 48) | A |
|  | Self-Evaluation of Negative Symptoms (SNS, 49) | S |
| Severity of positive symptoms | Scale for the Assessment of Positive Symptoms (SAPS, 50) | A |
| Interpersonal anhedonia | Anticipatory and Consummatory Interpersonal Pleasure Scale (ACIPS, 51) | S |
| Well-being | WHO-5 (52) | S |
| Depression | Calgary Depression Scale (CDSS, 53) | A |
| Cognitive assessment |  |  |
| Defeatist beliefs | Defeatist Performance Attitude Scale (DPAS, 54) | S |
| Readiness for therapy | Readiness for Therapy Questionnaire (RTQ, 55) | S |
| Facial emotion recognition | Emotion Recognition Task (ERT, 56) | T |
| Functional assessment |  |  |
| Daily life functioning | Personal and Social Performance Scale (PSP, 57) | A |
|  | Social Functioning Scale (SFS, 58) | S |
| Intervention-related assessment |  |  |
| VR immersion | Presence Questionnaire (59) | S |
| VR-induced motion sickness | Simulator Sickness Questionnaire (SSQ, 60) | S |
| Overall satisfaction | Client Satisfaction Questionnaire (CSQ, 61) | S |

*Note.* A, assessor-rated; S, self-reported; T, test.

#### Ecological Momentary Assessment (EMA)

Given their dynamic and fluctuating nature, capturing state NS is challenged by recall biases that may distort assessments based on retrospection (62). EMA allows for capturing transient mental states with high temporal resolution and has previously been used to assess NS (63–65), yielding improved ecological validity (66) and allowing for fine-grained analysis of subcomponents of NS, such as anticipatory and consummatory anhedonia (67). A specifically designed questionnaire will be issued to patients at five semi-random time points per day during two seven-day epochs at baseline (E1) and follow-up (E3). Participants will receive a notification on their phone, and the questionnaire will need to be completed within 15 minutes to be accepted. The questionnaire collects self-reported data concerning participants’ level of activity, affect, and company (see Appendix S4 for the complete EMA questionnaire and procedure). In between these high-frequency epochs (i.e., E1 and E3), a slightly altered version of the questionnaire will be issued each night, collecting the same data but referring to the entire previous day (E2).

#### Physical motion data

Sedentary behaviours (SB), which carry an increased risk for metabolic and cardiovascular diseases (68), are markedly more common in schizophrenia patients than in controls (69,70). NS have been found to be linked to higher levels of SB, and to attenuate the results of interventions aimed at increasing physical activity (71). Measurements relying on patients’ self-report are likely to underestimate SB (72). Given the complex interplay between physical inactivity and features of NS such as amotivation and avolition, this study will collect objective data on physical activity using the motion sensors of participants’ mobile phones and the study app. As a proxy of physical activity, the average daily step count will be assessed for E1 and E3 and compared between the two epochs.

#### Location data

As productive activities are more likely to happen away from home (73), this study will record participants’ geolocation as a functional outcome of changes in NS, using location data captured by the GPS sensors of participants’ mobile phones. The time spent within a 100-metre radius of the participants’ home address will be summed during E1 and E3 and compared between the two epochs.

#### Functional MRI

Both NS and motivational abnormalities reflect complex processes at a system level likely corresponding to functional brain activity within and between several regions and circuits (74). To elucidate neural correlates of reward anticipation and processing, we will conduct functional magnetic resonance imaging (fMRI) at baseline and follow-up using an adapted version of Knutsson et al.’s (75) monetary incentive delay task (MIDT) as described in Nielsen et al. (76), and supplementing it by a social incentive delay task (SIDT), in which faces displaying different emotions (77) are used as feedback stimuli rather than varying amounts of money (cf. 78). Comparing individuals with schizophrenia to healthy controls, meta-analysis of monetary reward-related blood oxygen level dependent (BOLD) signal changes has identified hypoactivation during reward anticipation in a network comprising the striatum, anterior and median cingulate cortex, amygdala, precentral gyrus, and superior temporal gyrus; while reward receipt was associated with hyperactivation of the striatum, insula, and amygdala, among others, and hypoactivation in the dorsolateral and medial prefrontal cortex (79). NS severity has been found to specifically correlate with reduced reactivity of the ventral striatum to social, but not monetary rewards (80). As the intervention targets social reward learning, we expect a normalisation of the neural response patterns to social rewards for participants in the intervention group.

#### Qualitative data

Given the complex nature of negative symptoms and the study’s aim of investigating feasibility and acceptability of a novel therapeutic intervention, we will additionally employ qualitative methods to shed light on aspects of patients’ and therapists’ experiences that may remain undiscovered by means of quantitative analysis (81,82). Semi-structured interviews with all patients in the intervention group will be conducted upon completion of the last therapy session, and a semi-structured group interview will be conducted with study therapists after all therapies have been finalised. Topics covered by the qualitative interviews will include the perceived usefulness, transferability, mechanisms of effect, and tolerability of the procedure. The qualitative investigation will also collect participants’ and therapists’ suggestions for improvement of the intervention.

### Data analysis

#### Statistical analysis

Feasibility and acceptability of trial procedures will be assessed by calculating proportions and 95% exact Clopper Pearson confidence intervals for recruitment, treatment retention, and satisfaction with therapy.

Signals of treatment efficacy will be explored by comparing secondary outcome measures between the control (TAU) and intervention (TAU + VRT) groups using analysis of covariance (ANCOVA), adjusting for baseline scores. EMA data will be analysed employing multilevel linear modelling (MLM).

#### Data management

The study, including all technical solutions, will comply with the General Data Protection Regulation (GDPR). Assessors conducting interviews with the participants will enter the data directly into an electronic case report form (CRF) using REDCap. When necessary, the data will be collected on paper and later entered in REDCap. Each participant’s data is connected to a unique serial number, and only assigned researchers have access to the data in REDCap. Questionnaires can be directly sent from REDCap to the secure public Danish mailing system (Digital Post) that is connected to each citizen’s personal identification number (CPR), and completed forms are returned digitally.

Data on paper, consent forms and other physical material with personal information are stored locally behind double locks. Imaging data and audio recordings will be saved on a logged network drive controlled by the Capital Region of Denmark, Centre for IT and Medical Technology (CIMT), granting only assigned researchers and therapists involved in the study access to the files.

### Ethics

The study has been approved by the National Committee on Health Research Ethics for the Capital Region of Denmark (H-24010871; date of final approval, 6 September, 2024). Informed consent will be obtained from all participants after they have been given oral and written information about the study. Participants are informed that participation is voluntary and that they can withdraw from the study at any time without adverse consequences to their treatment by the Mental Health Services. Side effects and adverse events will be monitored and documented throughout the study period, and any adverse events assumed to be related to the study will be reported to the Committee on Health Research Ethics of the Capital Region of Denmark. Aside from occasionally reported transient cybersickness (83), no adverse events are expected in direct relation to VRT.

### Study status

Recruitment of participants started 9 January, 2025, and is expected to end by 15 September, 2025. Data collection is projected to be completed by the end of December, 2025, and results are expected by the end of May, 2026.

## Discussion

Existing psychosocial treatments for SSD have demonstrated some efficacy in reducing NS, yet effect sizes remain generally small (9), and few interventions are specifically designed to target NS directly (22). Proposed mechanisms underlying NS include abnormalities in reward processing, dysfunctional beliefs, and impairments in social cognition – each offering pertinent targets for intervention (74). The present study aims to evaluate the feasibility and acceptability of a novel, symptom-specific, psychological intervention that specifically leverages social reward learning for reducing NS through VR technology, which has proven safe and efficacious in treating psychotic symptoms of psychosis (35).

This study has several strengths. First, it builds on a sound psychological rationale and integrates immersive VR to provide a realistic yet controlled environment for engaging the motivational system in ecologically valid scenarios. Second, it comprises a broad range of clinical, biological, behavioural, and qualitative outcomes, allowing for a multidimensional analysis of feasibility, acceptability, and signals of efficacy and mechanisms of change. Third, service users with lived experience were involved from the outset in shaping the intervention, enhancing its relevance and acceptability. Fourth, the controlled, assessor-blinded design of the study supports methodological rigor, creating robust groundwork for a potential follow-up efficacy trial.

A key limitation is the absence of an active control group. Instead, the intervention is compared to TAU without systematic assessment of its content or intensity. Given the heterogeneity of TAU, this may introduce variability that could affect outcomes (84). This limitation, however, appears acceptable within the context of a pilot study.

In sum, this pilot study will assess the feasibility and acceptability of a VR-based intervention addressing NS in patients with SSD. If successful, it will provide critical data to support the design of a fully powered randomised clinical trial and contribute to the development of more effective treatments for these persistently disabling symptoms.

## Data Availability

No datasets were generated or analysed during the current study. All relevant data from this study will be made available upon study completion.

## Conflicts of interest

The authors declare the following conflicts of interest: BE is part of the Advisory Board of Boehringer Ingelheim, Lundbeck Pharma A/S; and has received lecture fees from Boehringer Ingelheim, Otsuka Pharma Scandinavia AB, and Lundbeck Pharma A/S. Dr. Gregory Strauss is one of the original developers of the Brief Negative Symptom Scale (BNSS) and receives royalties and consultation fees from Medavante-ProPhase LLC in connection with commercial use of the BNSS and other professional activities; these fees are donated to the Brain and Behavior Research Foundation. Dr. Strauss has received honoraria and travel support from Medavante-ProPhase LLC for training pharmaceutical company raters on the BNSS. Dr. Strauss has consulted for and/or been on the speaker bureau for Minerva Neurosciences, Acadia, Lundbeck, Sunovion, Boehringer Ingelheim, Otsuka, and Johnson and Johnson pharmaceutical companies. LBG has received honorary from Lundbeck Pharma, Boehringer-Ingelheim, and Heka-VR.

## Notes

### Competing Interest Statement

I have read the journal's policy and the authors of this manuscript have the following competing interests: BE is part of the Advisory Board of Boehringer Ingelheim, Lundbeck Pharma A/S and has received lecture fees from Boehringer Ingelheim, Otsuka Pharma Scandinavia AB, and Lundbeck Pharma A/S. Dr. Gregory Strauss is one of the original developers of the Brief Negative Symptom Scale (BNSS) and receives royalties and consultation fees from Medavante-ProPhase LLC in connection with commercial use of the BNSS and other professional activities these fees are donated to the Brain and Behavior Research Foundation. Dr. Strauss has received honoraria and travel support from Medavante-ProPhase LLC for training pharmaceutical company raters on the BNSS. Dr. Strauss has consulted for and/or been on the speaker bureau for Minerva Neurosciences, Acadia, Lundbeck, Sunovion, Boehringer Ingelheim, Otsuka, and Johnson and Johnson pharmaceutical companies. LBG has received honorary from Lundbeck Pharma, Boehringer-Ingelheim, and Heka-VR.

### Clinical Trial

ClinicalTrials.gov registration ID: NCT06993831

### Funding Statement

Yes

### Author Declarations

The study is approved by the National Committee on Health Research Ethics for the Capital Region of Denmark (H-24010871). Informed consent will be obtained from all participants after they have been given oral and written information about the study.

